# Second monovalent SARS-CoV-2 mRNA booster restores Omicron-specific neutralizing activity in both nursing home residents and health care workers

**DOI:** 10.1101/2023.01.22.23284881

**Authors:** Clare Nugent, Yasin Abul, Elizabeth White, Fadi Shehadeh, Matthew Kaczynski, Lewis Oscar Felix, Narchonai Ganesan, Oladayo A. Oyebanji, Igor Vishnepolskiy, Elise M. Didion, Alexandra Paxitzis, Maegan L. Sheehan, Eleftherios Mylonakis, Brigid M. Wilson, Alejandro B. Balazs, Philip A. Chan, Christopher L. King, Walther M. Pfeifer, Evan Dickerson, David H. Canaday, Stefan Gravenstein

## Abstract

We examined whether the second monovalent SARS-CoV-2 mRNA booster increased antibody levels and their neutralizing activity to Omicron variants in nursing home residents (NH) residents and healthcare workers (HCW). We sampled 367 NH residents and 60 HCW after primary mRNA vaccination, first and second boosters, for antibody response and pseudovirus neutralization assay against SARS-CoV-2 wild-type (WT) (Wuhan-Hu-1) strain and Omicron BA1 variant. Antibody levels and neutralizing activity progressively increased with each booster but subsequently waned over weeks. NH residents, both those without and with prior infection, had a robust geometric mean fold rise (GMFR) of 10.2 (95% CI 5.1, 20.3) and 6.5 (95% CI 4.5, 9.3) respectively in Omicron-BA.1 subvariant specific neutralizing antibody levels following the second booster vaccination (p<0.001). These results support the ongoing efforts to ensure that both NH residents and HCW are up to date on recommended SARS-CoV-2 vaccine booster doses.

## Introduction

The Omicron SARS-CoV-2 variant and its sub-lineages including BA.1, BA.2, BA.2.12.1, BA.4 and BA.5 are more transmissible[1-3]. SARS-CoV-2 continues to impact nursing home (NH) residents disproportionately. To date, more than 1.3 million NH residents in the US have been infected with SARS-CoV-2 and over 160,000 have died [4]. Vaccination is the best intervention to reduce both mortality and morbidity in NH residents [5-7]. However, in the setting of new SARS-CoV-2 variants, there is still some uncertainty as to the optimal timing and frequency of consecutive booster vaccinations in this frail population.

In previous studies, six months following the two-dose BNT162b2 mRNA monovalent primary vaccine series, vaccine-induced antibody and neutralization titers in NH residents declined by more than 80%, and neutralization antibodies became undetectable in 57% of NH residents [8]. Interestingly, a first monovalent booster dose significantly enhanced vaccine-specific anti-spike and anti-receptor binding domain (RBD) antibody levels and neutralization activity above the pre-booster levels in NH residents [9]. In this study, we assessed anti-spike, anti-RBD antibody levels and neutralizing activity for Wuhan, Omicron BA.1 and BA.5 following a second monovalent booster in NH residents.

## Methods

### Ethical approval

This study was approved by the WCG Institutional Review Board. All participating individuals or their legally authorized representatives provided informed consent to be recruited into the study.

### Study Design and Study Population

The current analysis is part of an ongoing study [9, 10] in which NH residents and healthcare workers (HCWs) are consented and serially sampled before and after each SARS-CoV-2 vaccine dose. This analysis includes data collected between August 23, 2021, and September 8, 2022.

We recruited HCW and NH residents for this study. HCWs were individuals who worked at 4 NH buildings, 2 Cleveland area hospitals, and Case Western Reserve University; they had access to the same vaccine at the same time as the NH residents from 18 nursing homes in Ohio and 16 nursing homes in Rhode Island. Residents who received SARS-CoV-2 mRNA vaccines [(BNT162b2 (Pfizer-BioNTech) or mRNA-1273 (Moderna)] were included, and those who received the Ad26.COV2.S (Janssen) vaccine were excluded. Participants received their first booster dose 8-9 months after the primary vaccination series, and their second booster 4 to 6 months after the first booster. We report results from blood samples obtained at time points prior to and following the two booster doses. Participants were sampled at five time points: (1) pre-first booster (8-9 months post-primary series); (2) approximately 14 days post-first booster; (3) 4-6 months post-first booster; (4) approximately 14 days post-second booster; and (5) approximately 3 months post-second booster. Participants were included in the analysis if 2 or more included samples across these five time points were available.

### Definitions and Description of Data

### Prior or breakthrough Infection

We defined a participant as having had a prior SARS CoV-2 infection at the time of each vaccine dose based on (1) prior documentation in their medical chart of a positive polymerase chain reaction or antigen test; or (2) a significant increase in SARS-CoV-2 antibody levels that could not be explained by vaccination. Laboratory criteria supporting recent, or interval infection were a rise outside of lab variance of anti-spike, anti-RBD, nucleocapsid, or neutralizing assay results not accounted for by vaccination history.

### Infection Naive

We defined participants as “infection naive” at each time point if they did not meet the above criteria for prior infection. The prior infection/naive classification could change over time with infection history, i.e., individuals who had a SARS-CoV-2 infection between their first & second booster would be in the naive category for the first booster & ‘prior infection’ category for the second booster.

### Antispike and Anti-RBD Assay

We assessed vaccine-induced immune response using bead-multiplex immunoassay for anti-spike and RBD for SARS-CoV-2 wild-type (WT) (Wuhan-Hu-1) strain and Omicron BA1 variant as previously described [9]. Stabilized full-length spike protein (aa 16-1230, with furin site mutated), RBD (aa 319-541) from SARS-CoV-2 wild-type (WT) (Wuhan-Hu-1) strain and Omicron BA1 variant, and full-length N (aa1-419) from Wuhan were conjugated to magnetic microbeads (Luminex) and Magpix assay system (BioRad, Inc). Anti-Wuhan spike IgG levels were in Binding Antibody Units (BAU)/ml based on the Frederick National Laboratory standard, and anti-spike BA1 and anti-RBD from both strains are arbitrary units (AU)/ml.

### SARS-Cov-2 Pseudovirus Neutralization Assay

We produced lentiviral particles pseudotyped with spike protein based on the Wuhan and BA.1 and BA.4/BA.5 strains as previously described to define the neutralizing activity of vaccine recipients’ sera against coronaviruses [11]. We performed three-fold serial dilutions that ranged from 1:12 to 1:8748 and added to 50–250 infectious units of pseudovirus for 1 hour. pNT50 values were calculated by taking the inverse of the 50% inhibitory concentration value for all samples with a pseudovirus neutralization value of 80% or higher at the highest concentration of serum. The lower limit of detection (LLD) of this assay is 1:12 dilution.

### Statistical analysis

All analyses were performed in R version 4.2.2. Distributions of assays at the 5 time points were presented in boxplots. For all subjects with consecutive timepoints available, we assessed the geometric mean titer and its 95% confidence interval for all assays, strains, and time points, stratifying subjects at each booster by HCW vs. NH residents and infection naive vs. prior infection. We then calculated the geometric mean fold rise (GMFR) from pre to post boost for each booster dosing and performed t-tests on log-transformed titers comparing the observed fold rise to a null value of 1. All p values are shown without adjustment. Tests were not performed with 5 or fewer subjects.

## Results

The analytic sample that met criteria for inclusion included 427 volunteers including 367 NH residents (median age 75, 41% female, 78% white) and 60 HCW (median age 51, 57% female, 82% white) for comparison.

Prior data and the current study show a significant increase in anti-omicron titers from the prior time point to the post first boost that was similarly observed in this study.

There is then subsequent waning at 4-6 months after the first booster ranging between 70% to 95% for both binding antibody and neutralizing antibody titers. Most notable and robust declines of Omicron-specific anti-spike titers were from GMT 3771 to 509 AU/ml in NH residents with prior infection [geometric mean fold change (GMFC) of 0.13 (95% CI 0.09, 0.21)] (**Table 1**) and a decline of Wuhan specific anti-RBD levels from GMT 17041 to 300 AU/ml in NH residents with prior infection [GMFC of 0.02 (95% CI 0.01, 0.03)] (**Supplementary Table 1**). These types of declines provided rationale and support that prompted CDC recommendation for a second booster in older individuals.

**Table 1:**
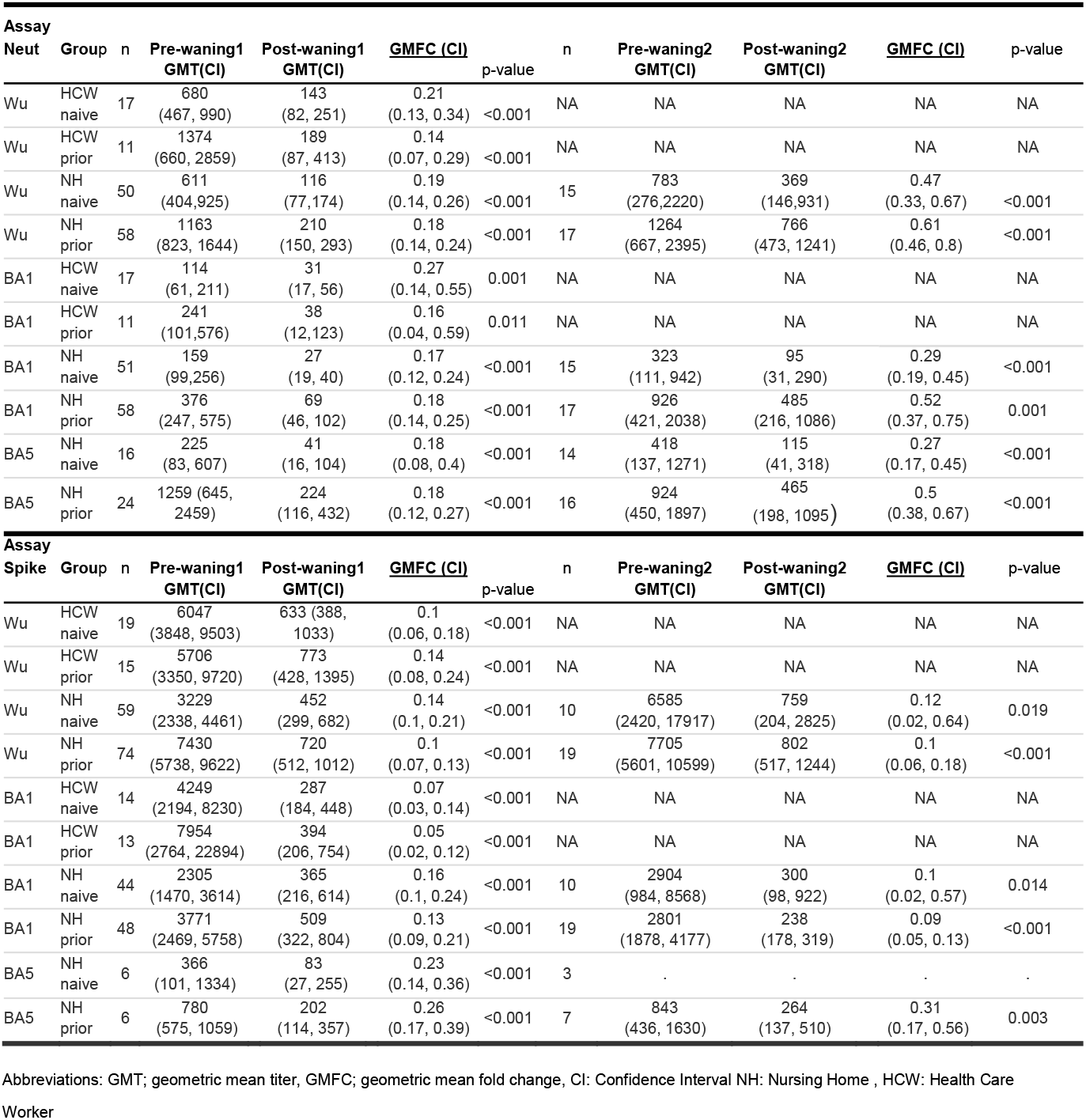
Waning of Wuhan and Omicron-specific antibody response after 4 to 6 months of first booster and second booster vaccination before the upcoming dose

NH residents, both those without and with prior infection, had a robust geometric mean fold rise (GMFR) of 10.2 (95% CI 5.1, 20.3) and 6.5 (95% CI 4.5, 9.3) respectively in Omicron-BA.1 subvariant specific neutralizing antibody levels following the second booster vaccination (p<0.001). Similarly, for infection-naive and prior infection their Omicron-BA.1 subvariant specific GMFR rose by 2.1 (95% CI 1, 4.2) and 3.4 (95% CI 2.4, 4.8) for anti-spike antibody (**Table 2**) and 2.4 (95% CI 1.1, 5.4) and 2.3 (95% CI 1.4, 3.9) for anti-RBD antibody levels, respectively (**Supplementary Table 2)**. We also observed a robust GMFR in Omicron-BA.5 subvariant specific neutralizing antibody levels [8.8 (95% CI 2.8, 27.8) and 6.9 (95% CI 4.4, 10.8) in each group, NH naive and NH prior] after second booster vaccination (**Figure 2 and Table 2**). Similar trends were observed in relatively smaller sample sizes of infection naive HCWs and HCWs with prior infection (**Table 2, Supplementary Table 2, Figure 1 and Supplementary Figure 1**) Subjects were then sampled 3 months after the second booster and, across all titers and groups, we also observed a notable decline in the geometric mean titers (GMT) from 2 weeks after the second booster to approximately 3 months after the second booster ranging between 50% to 95%. The most notable decline of Omicron-specific anti-spike titers were from GMT 2801 to 238 (AU)/ml in NH residents with prior infection [GMFC of 0.09 (95% CI 0.05, 0.13)] (**Table 1**) and another most notable decline was in Wuhan specific anti-Receptor binding domain levels from GMT 2109 to 167 (AU)/ml in NH residents with prior infection [GMFC of 0.08 (95% CI 0.05, 0.13)] (**Supplementary Table 1**).

**Table 2:** Neutralization and anti-spike titers against Wuhan and Omicron for NH residents and HCW with and without prior infection history

| Assay |  | Boost 1 GMT (CI) |  |  | Boost 1 GMFR |  | Boost 2 GMT (CI) |  |  | Boost 2 GMFR |  |
| --- | --- | --- | --- | --- | --- | --- | --- | --- | --- | --- | --- |
| Neut | Group | n | Pre | Post |  | p-value | n | Pre | Post |  | p-value |
| Wu | HCW naive | 23 | 14<br>(11, 17) | 804<br>(609, 1063) | 59.3<br>(45.6, 77.1) | <0.001 | 4 | . | . | . | . |
| Wu | HCW prior | 20 | 27<br>(14, 51) | 1287<br>(775, 2137) | 48<br>(27.9, 82.5) | <0.001 | 4 | . | . | . | . |
| Wu | NH naive | 41 | 13<br>(11, 16) | 509<br>(318, 815) | 38.1<br>(24.1, 60.3) | <0.001 | 31 | 170<br>(94, 307) | 1272<br>(746, 2170) | 7.5<br>(4.1, 13.8) | <0.001 |
| Wu | NH prior | 44 | 42<br>(25, 71) | 1094<br>(685, 1745) | 26<br>(15, 44) | <0.001 | 42 | 302<br>(202, 451) | 1195<br>(871, 1640) | 4<br>(3, 5.2) | <0.001 |
| BA1 | HCW naive | 23 | 14<br>(11, 18) | 143<br>(81, 254) | 10.2<br>(5.6, 18.5) | <0.001 | 4 | . | . | . | . |
| BA1 | HCW prior | 20 | 26<br>(15, 45) | 297<br>(168, 525) | 11.6<br>(6.2, 21.6) | <0.001 | 4 | . | . | . | . |
| BA1 | NH naive | 41 | 14<br>(11, 17) | 131<br>(75, 230) | 9.5<br>(5.6, 16.2) | <0.001 | 31 | 49<br>(24, 99) | 498<br>(253, 982) | 10.2<br>(5.1, 20.3) | <0.001 |
| BA1 | NH prior | 44 | 21<br>(15, 30) | 296<br>(170, 514) | 14<br>(8.1, 24.1) | <0.001 | 42 | 150<br>(88, 256) | 971<br>(664, 1420) | 6.5<br>(4.5, 9.3) | <0.001 |
| BA5 | NH naive | 3 | . | . | . | . | 18 | 91<br>(27,239) | 710<br>(249,2026) | 8.8<br>(2.8,27.8) | <0.001 |
| BA5 | NH prior | 12 | 46<br>(20, 108) | 815<br>(204, 3263) | 17.7<br>(4.6, 67.8) | <0.001 | 28 | 317<br>(182, 550) | 2189<br>(1477, 3245) | 6.9<br>(4.4, 10.8) | <0.001 |
| Assay |  | Boost 1 GMT (CI) |  |  | Boost 1 GMFR |  | Boost 2 GMT (CI) |  |  | Boost 2 GMFR |  |
| Spike | Group | n | Pre | Post |  | p-value | n | Pre | Post |  | p-value |
| Wu | HCW naive | 28 | 60<br>(36, 101) | 16961<br>(9659, 29781) | 62.1<br>(40.2, 95.9) | <0.001 | 4 | . | . | . | . |
| Wu | HCW prior | 25 | 147<br>(70, 309) | 21674<br>(11647, 40334) | 38.3<br>(20.2, 72.9) | <0.001 | 4 | . | . | . | . |
| Wu | NH naive | 90 | 13<br>(10, 17) | 4797<br>(2967, 7756) | 133.9<br>(95.8, 187.1) | <0.001 | 33 | 708<br>(418,1199) | 1637<br>(918, 2919) | 2.3<br>(1.1, 4.7) | 0.02 |
| Wu | NH prior | 126 | 315<br>(195, 508) | 13812<br>(9108, 20944) | 23.6<br>(15.7, 35.6) | <0.001 | 47 | 1370<br>(816,2298) | 5029<br>(3745, 6753) | 3.7<br>(2.6, 5.3) | <0.001 |
| BA1 | HCW naive | 17 | 67<br>(41, 109) | 3819<br>(2326, 6271) | 57.2<br>(32.6, 100.5) | <0.001 | 4 | . | . | . | . |
| BA1 | HCW prior | 18 | 235<br>(99, 557) | 13586<br>(6135, 30083) | 57.8<br>(23.7, 141.1) | <0.001 | 4 | . | . | . | . |
| BA1 | NH naive | 36 | 26<br>(13, 50) | 3131<br>(1875, 5227) | 120.5<br>(58.9, 246.4) | <0.001 | 33 | 332<br>(182, 607) | 683<br>(408,1142) | 2.1<br>(1, 4.2) | 0.046 |
| BA1 | NH prior | 35 | 216<br>(112, 417) | 5416<br>(3078,9532) | 25<br>(11.9, 52.5) | <0.001 | 47 | 534<br>(334, 856) | 1812<br>(1312, 2502) | 3.4<br>(2.4, 4.8) | <0.001 |
| BA5 | NH naive | 2 | . | . | . | . | 2 | . | . | . | . |
| BA5 | NH prior | 1 | . | . | . | . | 11 | 260<br>(132, 510) | 1195<br>(806, 1771) | 4.6<br>(2.9, 7.3) | <0.001 |

Abbreviations: GMT; geometric mean titer, GMFR; geometric mean fold rise, CI: Confidence Interval NH: Nursing Home , HCW: Health Care Worker

**Figure 1:**
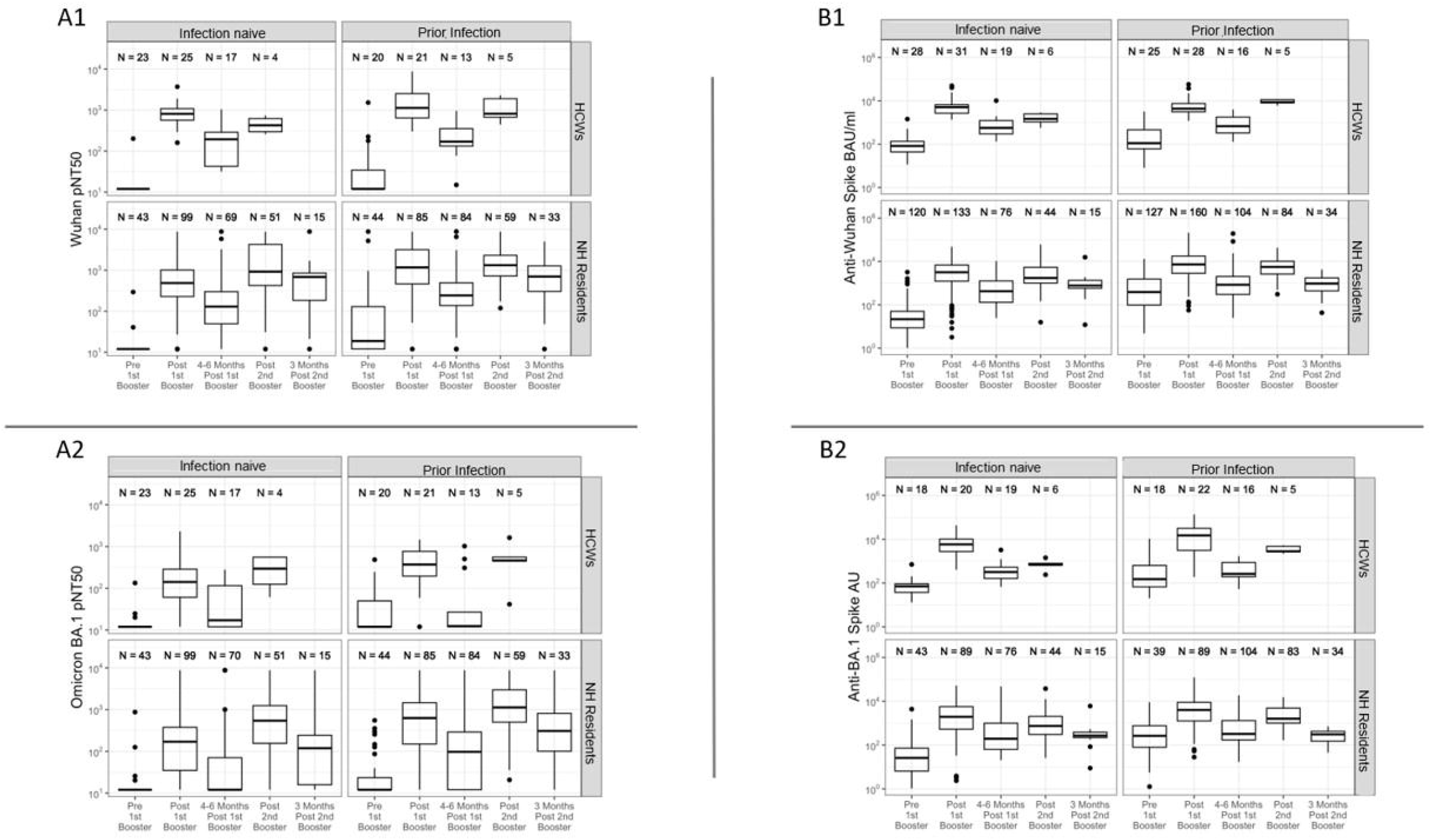
Neutralization titers (Panel A1:Wuhan ; A2: Omicron BA.1) and anti-spike levels (Panel B1:Wuhan; B2:Omicron BA.1) over time pre and post-boost with mRNA vaccination in healthcare workers (HCWs) and nursing home(NH) residents, with and without prior SARS-CoV-2 infection

**Figure 2:**
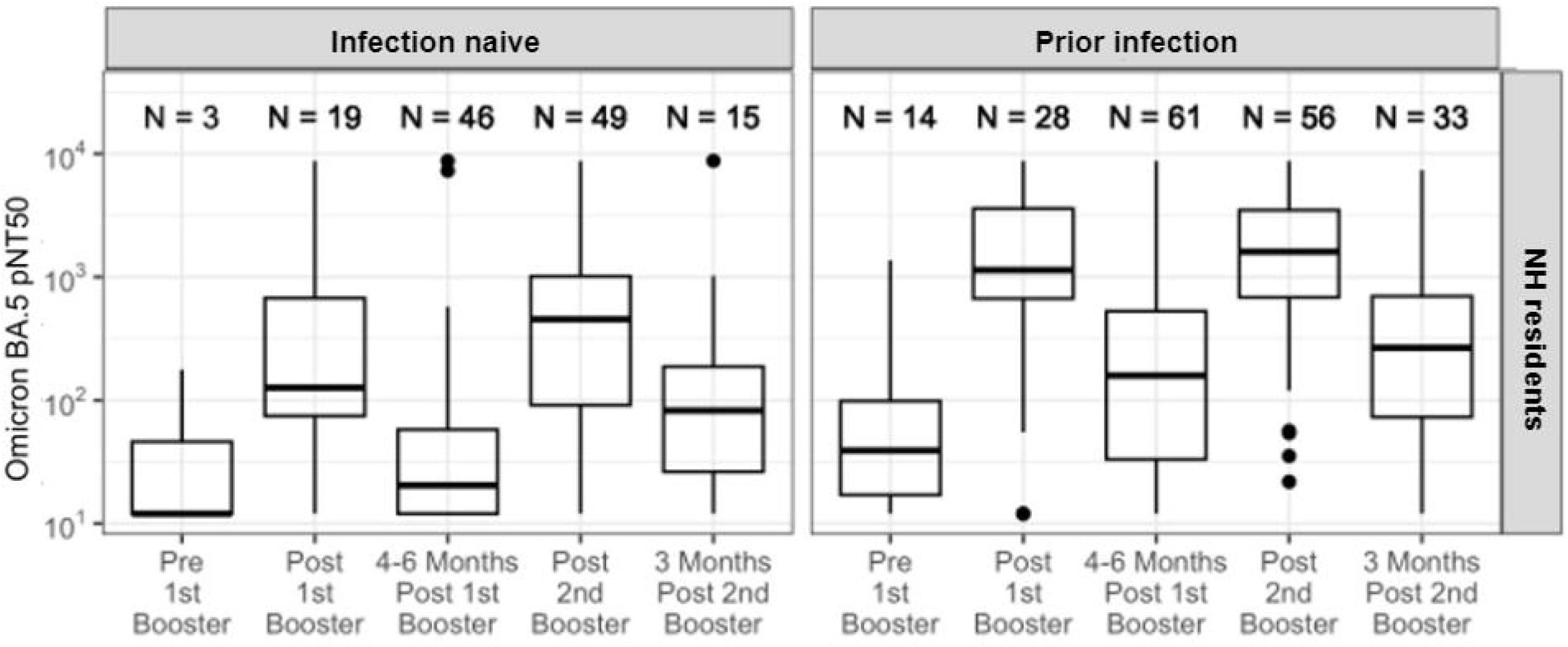
The rise and waning in pseudovirus neutralization(pNT50) antibody titers against Omicron BA.5 strain in naïve NH residents and NH residents with prior infection after 1st and 2nd boosters

## Discussion

NH resident anti-omicron (BA.1 and BA.4/BA.5) neutralization titers dropped substantially over the 6 months following the first mRNA monovalent booster. NH residents responded well to the second monovalent booster, achieving similar and in many cases higher levels than obtained after just one booster dose.

These findings are clinically important as neutralizing and binding antibody titers correlate with vaccine protection against many viral diseases [11, 12]. Anti-spike, anti-RBD and neutralization antibody titers correlate with enhanced protection from symptomatic SARS-CoV-2 infection [13, 14]. Feng et al reported that an anti-Wuhan spike level of 264 BAU/mL achieved 80% protection against symptomatic infection from the now ancestral alpha variant [14]. We found that all the NH residents in our sample achieved this cut off level after their second booster dose. However, Favresse et al proposed an alternative much higher cut-off of 8434 BAU/ml for binding antibodies to predict neutralization of the more recent Omicron BA.1 variant (sensitivity: 83.1%, specificity: 63.4%) [15]. The substantial difference in these two results demonstrates the complexity of defining precise antibody immune correlates in the current clinical setting where the evolving viral variants continue to acquire immune evasive mutations. Even as higher antibody titers may add protection, the relative clinical protection and titers decline over time with newer variants and will produce different thresholds of protection, perhaps especially in the frail older NH population in general and when compared to younger adults.

McConeghy et al showed that vaccine effectiveness of a second mRNA COVID-19 booster dose against SARS-CoV-2–associated hospitalization or death was 73.9% and 89.6% for death alone compared to a single booster dose among nursing home residents [6]. The mechanism for this increased protection is not yet fully defined but this current quantitative antibody response study after the second booster demonstrates that the additional doses for NH residents add to Omicron subvariant humoral immunity in this population. These higher titers likely play a significant role in this increased protection but certainly a contribution of cellular immunity which we did not measure may be a factor as well.

There were a few limitations of this study to note. First, was the relatively small sample size. Second, we were unable to obtain follow up samples from all participants. Third, while women make up over ⅔ of US NH residents, our sample of 41% women due to the large contribution of Veterans in state Veterans homes, our sample skews from that of the US nursing home population. Fourth, we did not assess vaccine induced T-cell immunity. Fifth, we may have misclassified some participants with undetected asymptomatic SARS CoV-2 infection who did not meet the laboratory criteria threshold for prior infection between infection naive and prior infected groups. The HCW sample size was small, hampered also by those under 50 who were not eligible for a second booster per CDC guidelines. The HCW remaining for analysis therefore skewed towards better representing older individuals in this group. Finally, as the subjects recruited after prior SARS-CoV-2 infection all survived the infection, their inclusion may introduce survivorship to our analysis.

## Conclusion

In a sample of 367 nursing home residents, anti-Omicron neutralization titers waned significantly over the months following the first and second SARS-CoV-2 mRNA monovalent booster doses. Antibody titers rose significantly with boosting, achieving similar or in many cases higher levels after the second boost than after the first, while also gaining important anti-Omicron neutralizing activity. Subsequent waning of antibody titers with additional boosters and broadening of neutralizing immunity with the second boost provides evidence of the critical need to keep nursing home residents, a particularly vulnerable population, up to date with SARS-CoV-2 vaccination.

## Data Availability

All data produced in the present study are available upon reasonable request to the authors

## Grant support/Acknowledgement

This work was supported by NIH AI129709-03S1, CDC 200-2016-91773, U01 CA260539- 01, and VA BX005507-01

## Figure Legends and Tables

**Supplementary Table 1:** Waning of anti-Receptor binding domain levels after 4 to 6 months of first booster vaccination before the second booster

| Assay |  | n | Pre-waning1<br>GMT(CI) | Post-waning1<br>GMT(CI) | GMFC (CI) |  | n | Pre-waning2<br>GMT(CI) | Post-waning2<br>GMT(CI) | GMFC (CI) | p-value |
| --- | --- | --- | --- | --- | --- | --- | --- | --- | --- | --- | --- |
| RBD | Group |  |  |  |  | p-value |  |  |  |  |  |
| Wu | HCW<br>naive | 19 | 21773<br>(12580, 37683) | 496<br>(216, 1143) | 0.02<br>(0.01, 0.06) | <0.001 | NA | NA | NA | NA | NA |
| Wu | HCW<br>prior | 15 | 152687<br>(11993, 57615) | 540<br>(193, 1510) | 0.02<br>(0.01, 0.05) | <0.001 | NA | NA | NA | NA | NA |
| Wu | NH<br>naive | 59 | 4211<br>(2353, 7537) | 147<br>(80, 271) | 0.03<br>(0.02, 0.07) | <0.001 | 10 | 2537<br>(925, 6963) | 204<br>(123, 339) | 0.08<br>(0.02, 0.28) | 0.001 |
| Wu | NH<br>prior | 73 | 17041<br>(10164, 28571) | 300<br>(182, 493) | 0.02<br>(0.01, 0.03) | <0.001 | 19 | 2109<br>(1303, 3413) | 167<br>(115, 240) | 0.08<br>(0.05, 0.13) | <0.001 |
| BA1 | HCW<br>naive | 14 | 1169<br>(480, 2851) | 280<br>(155, 506) | 0.24<br>(0.09, 0.63) | 0.007 | NA | NA | NA | NA | NA |
| BA1 | HCW<br>prior | 13 | 931<br>(315, 2757) | 205<br>(103, 407) | 0.22<br>(0.11, 0.45) | <0.001 | NA | NA | NA | NA | NA |
| BA1 | NH<br>naive | 42 | 623<br>(331, 1171) | 212<br>(102, 439) | 0.34<br>(0.2, 0.59) | <0.001 | 10 | 4945<br>(814, 30034) | 420<br>(116, 1521) | 0.08<br>(0.02, 0.47) | 0.01 |
| BA1 | NH<br>prior | 46 | 1253<br>(793, 1981) | 450<br>(232, 872) | 0.36<br>(0.21, 0.62) | <0.001 | 19 | 4647<br>(2483, 8696) | 292<br>(195, 438) | 0.06<br>(0.03, 0.12) | <0.001 |
| BA5 | NH<br>naive | 6 | 294<br>(46, 1885) | 66<br>(13, 326) | 0.22<br>(0.12, 0.41) | 0.001 | 3 | . | . | . | . |
| BA5 | NH<br>prior | 6 | 366<br>(240, 557) | 88<br>(39, 197) | 0.24<br>(0.15, 0.39) | <0.001 | 7 | 555<br>(249, 1239) | 177<br>(100, 314) | 0.32<br>(0.16, 0.63) | 0.006 |

Abbreviations: GMT; geometric mean titer, GMFC; geometric mean fold change, CI: Confidence Interval NH: Nursing Home , HCW: Health Care Workers

**Supplementary Table 2:**
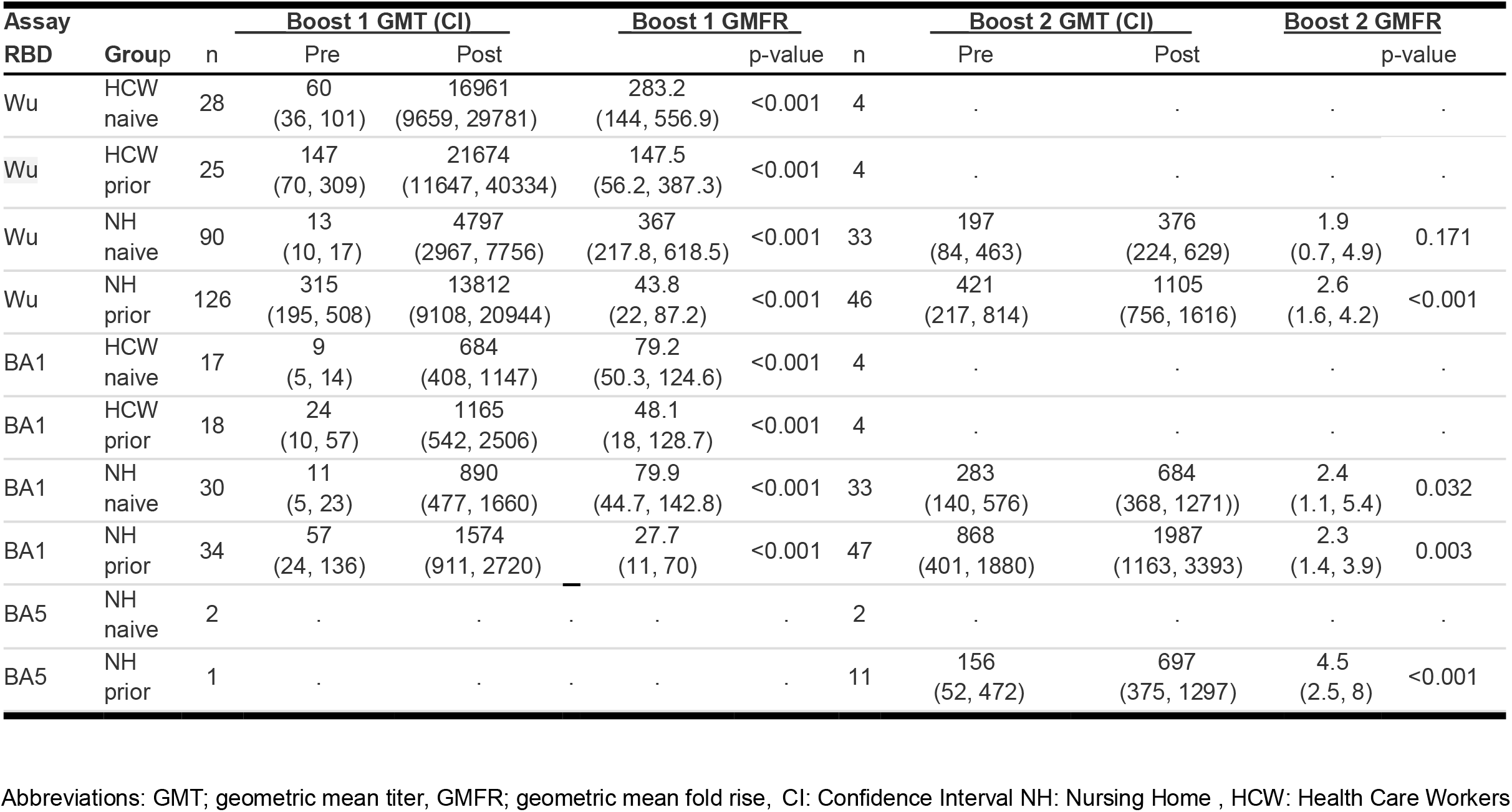
Anti-Receptor binding domain levels against Wuhan and Omicron for NH residents and HCW with and without prior infection history

