## Supplementary figures and images for "Second monovalent SARS-CoV-2 mRNA booster restores Omicron-specific neutralizing activity in both nursing home residents and health care workers"

### Supplemental Figure 1

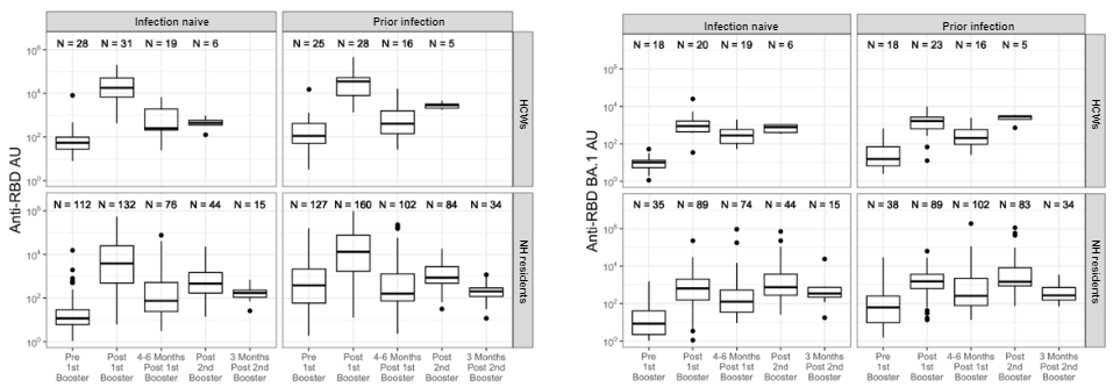
